# Laboratory capacity assessment in a resource-limited health system, Savannah Region, Ghana; a descriptive cross-sectional study

**DOI:** 10.64898/2026.04.08.26350443

**Authors:** Farouk U. Saeed, Chrysantus Kubio, Richard Kutame, Gifty Boateng

## Abstract

**Background:** Laboratory services are essential to the provision of health service delivery across the world. In resource-constrained settings such as in low- and middle-income countries like Ghana, maintenance of a strong capacity could be more challenging. This study assessed the capacity and gaps in laboratory service delivery in three districts of the Savannah Region of Ghana.

**Methods:** The WHO laboratory assessment tool (LAT) was adapted to collect data in 10 health facilities based on 11 operational system modules. Data were collected through interviews. Capacity was defined based on a 100-point score scale and interpreted as weak (<50%), moderate (50-80%) and strong (>80%). Differences in median scores were determined using Friedman and Kruska-Wallis tests. Statistical significance was set at p < 0.05. A scale (0-5) was used to identify the needs of the laboratory.

**Results:** Overall, capacity score was moderate, mean 50% ± 25.7 with a median score of 52.5%, IQR: 30.0-68.5%. Testing module received the highest score, 71.5%, while document module scored the lowest, 14.5%. Scores did not differ significantly between system components after multiple comparisons, p>adjusted alpha. Hospital-level laboratories performed significantly higher than polyclinics (adjusted p = 0.044) and health centers (adjusted p<0.001). The biggest needs were biosafety, equipment maintenance, human and financial resources (median gap score: 3-4).

**Conclusion:** The laboratory capacity in the health system of the Savannah Region was moderate, requiring improvements in all operational areas. The biggest needs include safety, equipment, human and financial support systems. Addressing these critical gaps would have direct impact on public health and patient outcomes.

## Introduction

Globally, medical laboratory services contribute to the majority of medical decisions. In the United States of America, more than 70% of medical decisions are hinged on results from the laboratory [1]. Laboratory services are essential and critical to the provision of health service delivery across the world. Laboratories offer accurate, reliable and timely diagnostic services for the prevention, treatment, cure and management of diseases and conditions critical to patient care. As new technologies and evidence-based practices emerge, laboratories have become the backbone to modern medicine and public health [2].

The burden of public health priority diseases, such as non-communicable diseases (NCDs) and infectious diseases, continues to increase in Africa [3]. These diseases can have similar or overlapping signs and symptoms, but laboratory diagnosis provides accurate and reliable information for better patient outcomes. Therefore, maintenance of a strong laboratory capacity is crucial. In situations where this may be lacking, the consequences can lead to a missed opportunity to detect an outbreak which could spread to neighbouring countries. Moreover, the World Health Organization (WHO) recommends its member states to strengthen national surveillance and response as part of the core International Health Regulation (IHR) capacities [4]. One such core requisite of the IHR is the capacity to detect an outbreak early through strengthened laboratory systems [5]. In resource-constraint settings, such as in low- and middle-income countries (LMICs) like Ghana, maintenance of such capacity could be more challenging [6].

Ghana’s health system is organized into various tiers referred to as the levels of care [7]. These levels of care depict how health care is provided in the communities, districts and regions. At the base of this level of care is the primary healthcare (PHC) system, which is responsible for providing general preventive and curative services. The primary care providers are the first point of contact for most people in the community. Above the PHC are the secondary and tertiary system. Laboratory services are organized along these lines but with different focus areas. Public health, clinical and veterinary laboratories operate with different client bases [8]. Since laboratories operate in different systems and networks, it is important to assess the capacity at the different levels of health service delivery to understand their readiness and capabilities.

The WHO laboratory assessment tool (WHO LAT) offers laboratories in low-resource countries the opportunity to assess their capacities and readiness to provide accurate, reliable and timely laboratory services [9]. Since its development, the tool has been used to assess different laboratories for different purposes [10]–[12]. In China, the tool was used to assess capacity for fungi testing in a low-resource setting [11]. Markby et al. assessed laboratory capacities in two conflict-affected regions of Gaza and DRC [13]. In Ghana, the tool has been used to assess the capacity of the national public health laboratory for molecular testing [12]. However, to the best of our knowledge, no study has been conducted in the Savannah Region to assess the capacity of laboratories. Assessing the capacity of laboratories will provide objective information for decision makers to implement capacity-strengthening activities. The study assessed the capacities and gaps in service delivery of medical laboratories in three districts of the Savannah Region.

## Methods

### Study design

A descriptive cross-sectional study design was employed to conduct the study between 1^st^ to 31^st^ March 2025.

### Study area

The study was conducted in the Savannah Region, which occupies about 15% of the total land mass of Ghana. The region has a population of 653,266 with an annual growth rate of 2.1% and a population density of 18.8 persons per square kilometer (km^2^) [14]. The region shares international boundaries with La Cote D’Ivoire and Burkina Faso to the West. Health service in the region is provided by a network of district and sub-district health facilities **(Fig 1)**. The region has seven administrative health districts and forty-two sub-districts. The region falls in the meningitis belt of Ghana and outbreaks of meningitis are not uncommon [15]. In the recent past, outbreaks of Marburg Virus Disease and Yellow Fever have been recorded in the region [16], [17].

**Fig 1.**
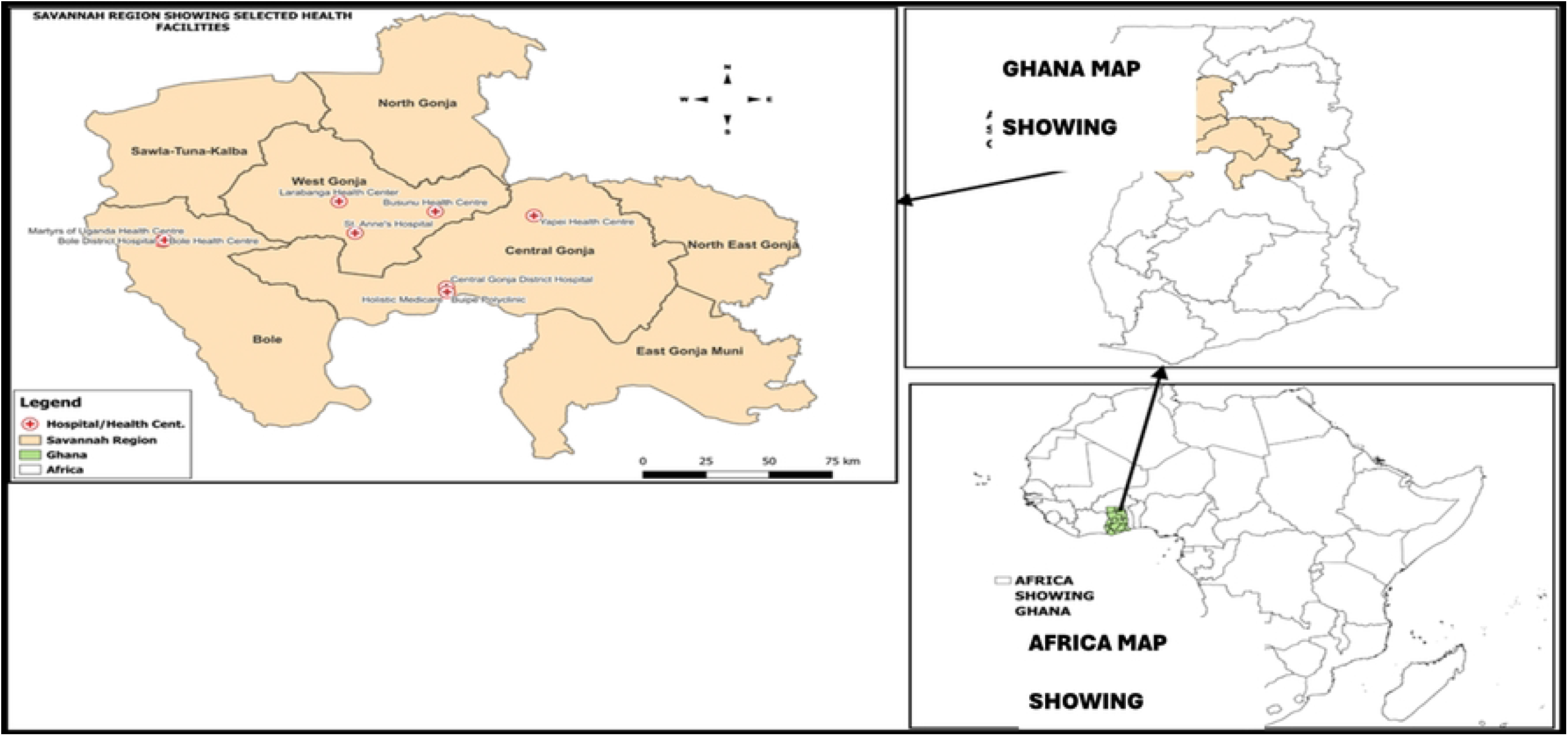

### Sample size and selection of sample

The sample size was determined based on the knowledge of the different management, funding mechanisms and the levels of the health delivery system that exist in the health sector of the region. Due to the poor road network in the region, access to some facilities was a challenge. As a result, a stratified convenient sampling technique was employed to select a mix of four hospitals (57% of all hospitals), three polyclinics (50% of all polyclinics) and three health centers (38% of all health centers) with functional laboratories. These facilities were selected from three districts and included private-for-profit, public, and faith-based ownerships.

### Description of the assessment tool

The assessment tool used was the WHO LAT (version 2012) facility level questionnaire. The WHO LAT is a generic document used to assess laboratory capacity in resource limited settings. It provides a standardized and objective assessment of laboratories for decision makers to plan and implement laboratory strengthening activities. The tool consists of eleven (11) modules each assessing different functionalities of the laboratory system. The modules are namely: organization and management; document; specimen collection, handling and transport; data and information management; consumables and reagent; equipment; testing; human resources; facilities; biorisk management and public health functions. Each module has a set of questions that address each area of laboratory operations. The questions have been assigned marks with every response giving a score between 0-100%. A “Yes” response is assigned 100 percentage points, a “Partial” is assigned 50% and a “No” is assigned a 0%. Questions that are not applicable do not lead to a score and do not contribute to the overall score. The final assessment score is calculated as the average score of the eleven modules assessment scores. The tool is also used to collect information on laboratory identification, but this module is non-scoring.

### Source of Data

Data was obtained through interviews with laboratory managers, public health officers and clinicians in the various health facilities and direct observations of laboratory work practices. Data on diagnostic testing was collected from laboratory registers and the laboratory information management system (LIMS) database. Quality and technical records were reviewed and assessed.

### Data collection

Data was collected using the WHO LAT (version 2012) facility questionnaire. The tool was adapted to suit the local situation and requirements (i.e. non-applicable areas in the organization and management, public health modules were withdrawn). A team of data collectors were trained on the use of the data collection tool. A pretest of the questionnaire was done, in laboratories that were not part of those assessed, to ensure quality data collection. The data collection officers were trained and experienced medical laboratory scientists. The tool was sent to the laboratory managers through a WhatsApp platform before the visits to ensure respondents understood the tool. This was followed by a field visit to administer the questionnaire. After the data collection, immediate feedback was given to the laboratories to help them address the areas that needed improvement.

### Data analysis

The data were entered into Microsoft Excel (version 19) and exported into SPSS (version 21, IBM) for analysis. We double-checked the data and reconciled where there were inconsistencies. Descriptive statistic was used to present ordinal data as means and medians with interquartile ranges (IQR). Laboratory capacity was defined based on a 100-point scale with scores <50% interpreted as weak, moderate (50-80%), and strong (>80%). A gap analysis was conducted using data collected in certain key areas of laboratory operations to identify strengths and weaknesses. A scale between 0 and 5 was used to identify the biggest needs of the laboratory, with 0 being no gap and 5 being the biggest gap. Differences in median scores were analysed using Friedman test, a non-parametric test in SPSS suitable for ordinal data for matched pairs. Statistical significance was set at p < 0.05. A post-hoc analysis was done with multiple pairwise comparisons for significant tests using Holm-Bonferroni adjustments. To compare differences in medians between the laboratory types, a Kruskal-Wallis H test was done using the non-parametric function in SPSS. The results were presented as tables and charts.

## Results

### Demographic characteristics of laboratories

Out of ten (10) health facilities that were selected to participate in this study, seven (7) were peripheral level laboratories and three (3) were intermediate levels. Of the four hospitals that were selected, four hospitals, three polyclinics and three health centers were selected. Majority were public-owned facilities (80%). All the laboratories were affiliations of the human sector **(Table 1)**.

**Table 1.** Characteristics of laboratories in the WHO LAT assessment, Savannah Region, Ghana, 2025.

| Characteristic | Frequency, N=10 (%) |
| --- | --- |
| **Level of Laboratory** |  |
| Central | 0 (0%) |
| Intermediate | 3 (30%) |
| Peripheral | 7 (70%) |
| **Type of Laboratory** |  |
| Hospital | 4 (40%) |
| Polyclinic | 3 (30%) |
| Health Center | 3 (30%) |
| **Ownership of Laboratory** |  |
| Public | 8 (80%) |
| Private | 1 (10%) |
| Faith-based | 1 (10%) |
| **Affiliation of Laboratory** |  |
| Health | 10 (100%) |
| Animal | 0 (0%) |

### Performance of the laboratories in the WHO LAT assessment

Overall, the facilities in this assessment received a moderate mean score, 50% ± 25.7 SD with a median score of 52.5%, IQR: 30.0-68.5%. The laboratory component indicator with the highest and least median scores were laboratory testing performance (71.5%) and documents (25.4%) respectively. The laboratories also received moderate median scores in specimen collection, handling and transport (69.0%), facilities (67.5%), consumables and reagents (52.5%), data and information management (57.0%), equipment (53.0%) and human resource (50.0%). In the other modules, the laboratories received a weak median score, which included organization and management (32.0%), public health functions (30.5%) and biorisk management (21.5%) **(Fig 2)**.

**Fig 2.**
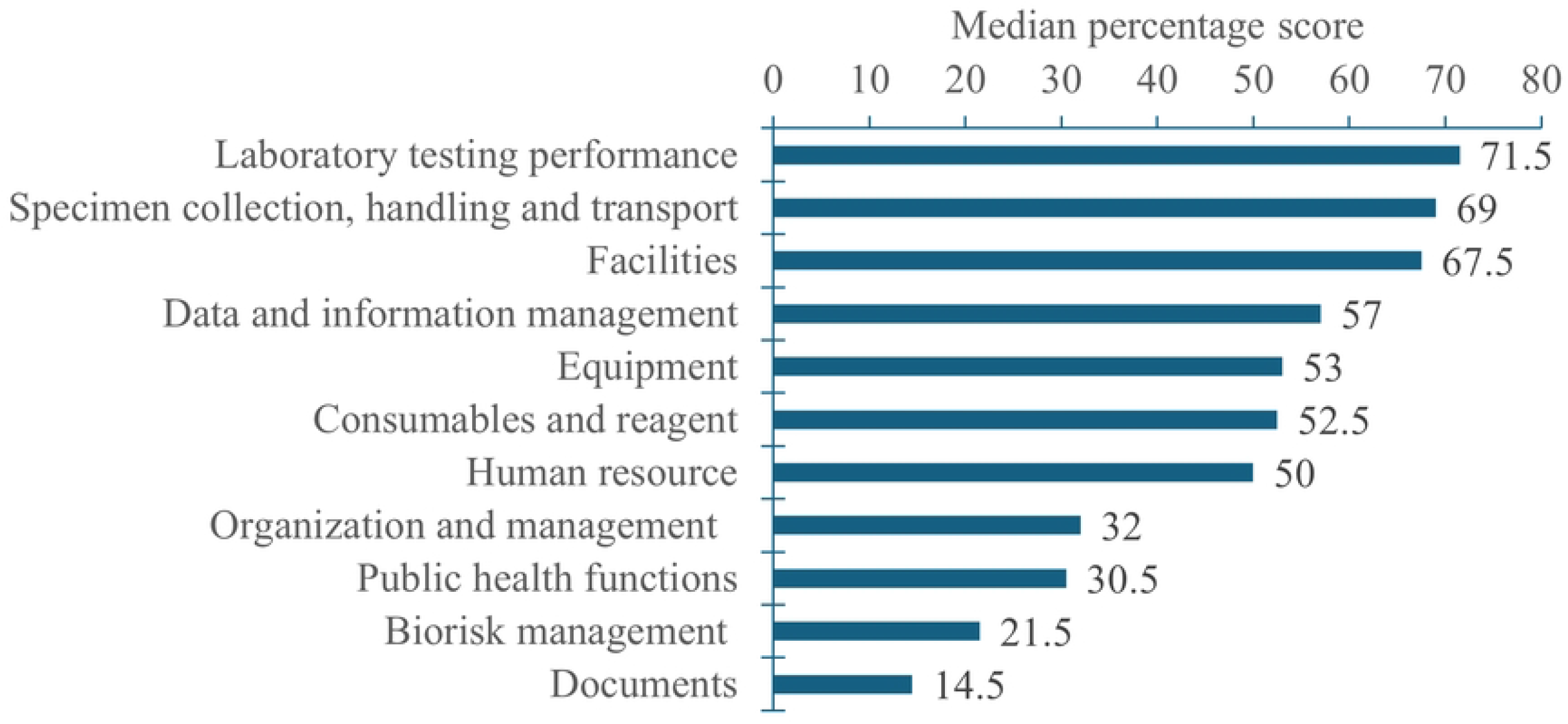

### Comparison of laboratory module indicators’ performance

The Friedman test indicated statistically significant difference in median percentage scores across laboratory module indicators (Friedman chi-square (10) = 60.78, p < 0.001). However, post-hoc pairwise comparison using Wilcoxon signed-rank test with Holm-Bonferroni corrections, adjusted alpha=0.0009, revealed no statistically significant difference between any of the component indicators.

### Comparison of assessment score by laboratory type

Median score for hospital laboratories was moderate, 62.5%, IQR:54.5-83.5. Polyclinic and health center level laboratories received weak median scores, 49.0% IQR=27.0-63.0 and 34.0% IQR=21.0-42.0% respectively. The median laboratory assessment scores differed significantly between laboratory types (Kruskal-Wallis chi-square (2) = 25.4, p<0.001). Post-hoc analysis with Bonferroni adjustment indicated that hospitals performed better than polyclinics (adjusted p = 0.044) and health centers (adjusted p<0.001) whereas polyclinics performed better against health centers (adjusted p=0.046) **(Table 2)**.

**Table 2.** Median assessment score by laboratory type in the Savannah Region, Ghana, 2025.

| No. | Laboratory type | N | Median (%) | IQR (%) |
| --- | --- | --- | --- | --- |
| 1 | Heath Center | 33 | 34.0 | 21.0-42.0 |
| 2 | Hospital | 44 | 62.5 | 54.5-83.5 |
| 3 | Polyclinic | 33 | 49.0 | 27.0-63.0 |

### Findings from the Gap Analysis

The gap analysis showed that the scale of needs among laboratories varied with the biggest needs in human resource qualification and availability, equipment calibration and maintenance, laboratory safety and security, and financial resource (median score = 3-4) **(Fig 3)**. The Friedman test of median scores indicated significant difference in the scale of needs among laboratories (Chi-square (12) = 42.8, p < 0.001) with a fair agreement among the raters (Kendall W = 0.36, p<0.001). After pairwise comparison with Holm-Bonferroni corrections, adjusted p = 0.0006, there was no significant difference between the needs of laboratories.

**Fig 3.**
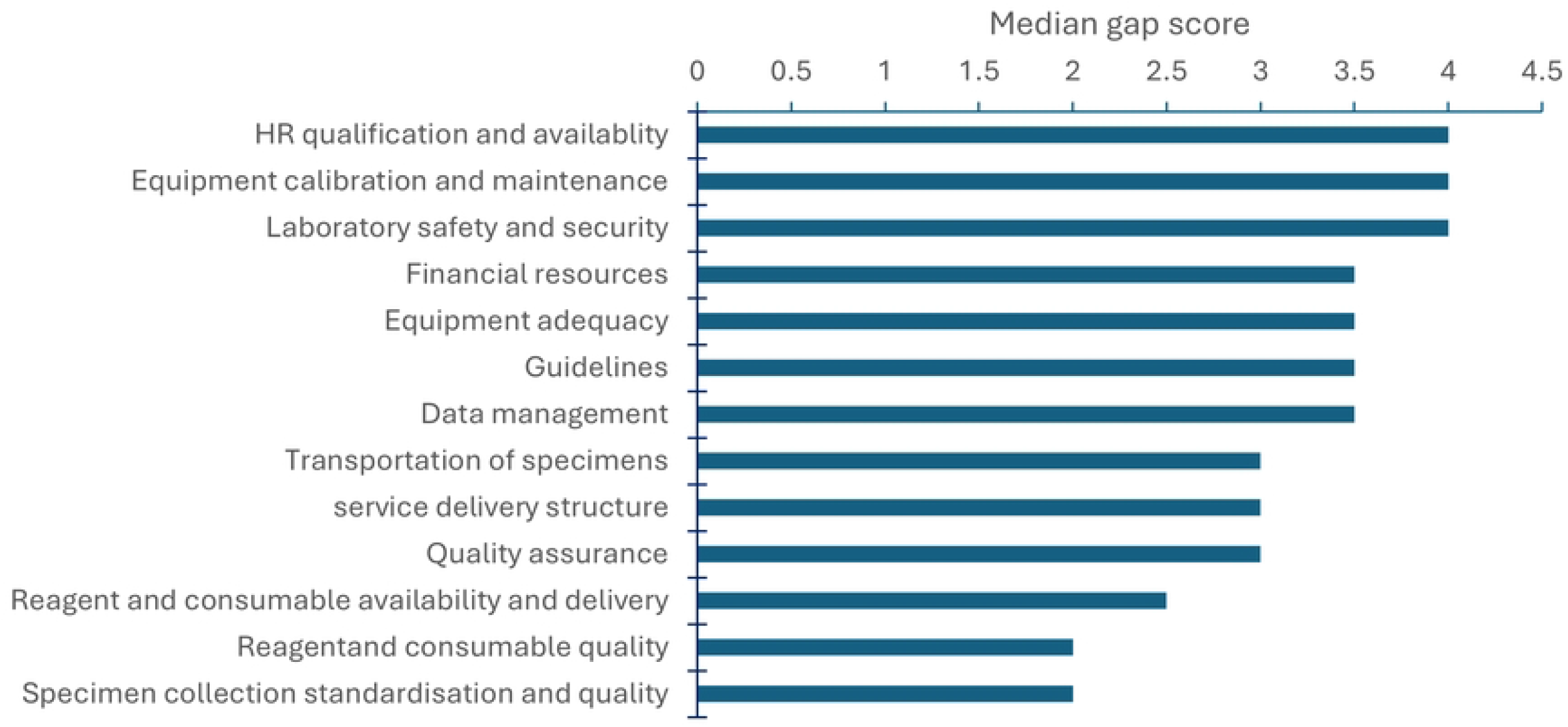

### Testing Disciplines

Majority of tests done per month were point-of-care testing (POCT) (40.2%). Approximately 28.8% were parasitology tests and about one-third (26.8%) of test were haematology and haemostasis. Small proportions were recorded in disciplines like clinical chemistry (1.8%), transfusion blood testing (1.6%), and molecular testing (0.8%) **(Fig 4)**.

**Fig 4.**
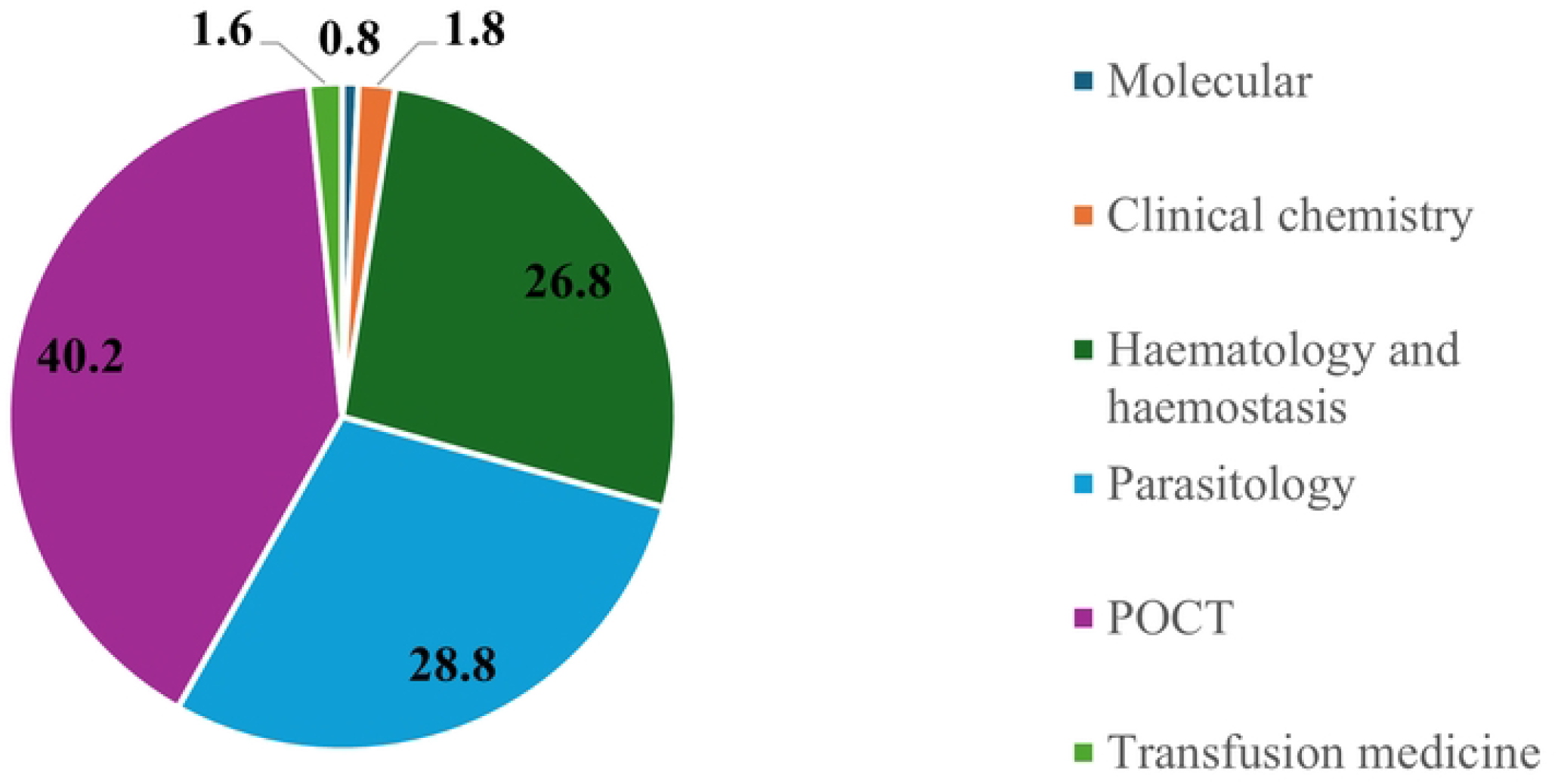

## Discussion

The study was conducted to assess the capacities of laboratories in resource-constrained districts of the Savannah Region. The laboratories in this assessment received an overall moderate score indicating inadequate capacity requiring some improvements particularly in areas such as documents and biorisk management. Even though scores differed between indicators, it was not statistically significant. Among the types of laboratories, hospital-level performed significantly better than their lower-tiered counterparts. The laboratories expressed varied needs with the biggest needs in areas such as HR, equipment, biosafety and financial resources. The most common testing discipline in the region was POCT followed by parasitological tests.

Overall, the laboratories in this assessment received moderate score indicating that although there is evidence of essential system functions, there remain important areas that require strengthening. This finding has been corroborated by a similar finding by Markby J. et al. in their study of laboratory capacity in the conflict afflicted Democratic Republic of Congo [10]. Moderate performance levels are commonly reported in health system laboratory assessments in LMICs where constraints such as inadequate infrastructure and shortages of trained personnel may affect the full implementation of laboratory quality management systems. This could have serious implications for both clinical and public health outcomes if some interventions are not implemented to improve the current status.

Substantial differences were observed in the different assessment modules with document and biorisk management modules underperforming while test performance was moderate. This suggests variations in the level of implementation of laboratory components across system domains. Variability among laboratory components is not uncommon, as different system components often receive varying levels of attention, resources and technical support. For instance, managers with constrained resources may choose to invest in components such as testing performance, specimen collection and facilities as opposed to biorisk and documents due to the complexities and financial burden of implementation of quality management systems. This finding is similar to a study in Ethiopia [13] on laboratory capacity assessment. Quality management programs are critical for efficient laboratory service delivery [18]. Moreover, it was observed during the assessment that the laboratories in the region have a cultural preference for verbal communication. Some laboratories have developed the habit of performing procedures without necessary documentation. This practice may contribute to the low score in the document module. Even though performance differed between modules, statistical significance was not achieved. This could be as a result of the strict statistical corrections and small number of laboratories that were applied. Nonetheless, the results emphasize the need for managers to provide interventions in all aspects of the laboratory operations.

In the context of laboratories levels used in this assessment, the analysis showed significant differences among the levels suggesting variations in the performances at the different levels of care. Hospital-level laboratories performed significantly better against their lower-tiered counterparts. The comparison has emphasized a strong funding gap in the laboratories in relation to equipment adequacy, recruitment of qualified and adequate staff, laboratory safety, implementation of quality management system and laboratory public health surveillance. The health authorities in Ghana recently introduced a data management system in selected health facilities across the country. This could have contributed a lot to the performance of the hospital and polyclinic level laboratories in data management. While this may be a prudent management decision, it could lead to undermining efficiency and quality service delivery at the lower levels. Perhaps, it is an indication that health authorities in the region channel their resources towards the highest level within the primary health care setting. But the situation at the health center level is not promising and requires urgent attention particularly because they are the first point of contact for most patients that need laboratory service.

Laboratories differed significantly in their scale of needs with the major gaps identified in biosafety, equipment calibration and maintenance, human and financial support systems. Conversely, minor gap scores were observed in reagent quality, and specimen collection practices. While overall variability exists in gap scores, statistical significance was not attained after adjustment for multiple comparisons. Consequently, strengthening laboratory systems require comprehensive system approach that addresses multiple components concurrently rather than focusing on isolated areas of improvement. Managers of laboratories in this assessment considered these gaps as their biggest priorities but this is not a peculiar trend in healthcare systems with resource challenges.

Although POCT is not a discipline of laboratory science, it has evolved in the health space for screening purposes owing to the demand for rapid test results for patient management [19]. Its use covers areas such as antigen and antibody screening for HIV, malaria parasite, syphilis, hepatitis B and C, and some hormones. Other uses include haemoglobin and glucose measurements. Disciplines such as bacteriological culture and sensitivity, cytology, water, food and veterinary tests were not available. Laboratories in the region are part of a network of laboratories in the country with different referral mechanisms [20]. This network helps the laboratories to transport samples to a higher-level facility where testing can be done. Despite this intervention, the testing capacity is still low compared to the national recommendation for testing at the district and sub-district levels [21]. This has also re-emphasized the need for improved funding to improve capacity, particularly for the health center laboratories, due to the long travel distances to the referral laboratories.

## Limitations of the Study

Despite the useful insights into the performance of laboratory systems, the study has several limitations that should be considered when interpreting the findings. Firstly, the relatively small number of health facilities may have limited the statistical power to detect significant differences between specific laboratory systems during post-hoc analysis. Consequently, our findings cannot be appropriately used to represent the entire situation in the region. However, the differences that may occur could be minimal. Secondly, although standardized assessment procedures were used, variations in documentation practices across facilities could have influenced the scoring of certain components. Thirdly, the study being a cross-sectional design cannot account for potential temporal changes in laboratory capacity. Lastly, even though the capacities of laboratories were assessed, it does not intend to describe in a perfect way the laboratory system in the Savannah Region.

## Conclusion

The laboratory capacity in the Savannah Region of Ghana is moderate. Performance varied among system components with no significant differences, though, hospital-level laboratories performed better than their lower-tiered counterparts. The greatest weaknesses were documentation, biosecurity and biosafety, equipment calibration and maintenance, human and financial support systems. These gaps have direct implications for public health response, especially in times of outbreak or routine disease monitoring. Addressing these challenges will require the health authorities to invest in capacity-building programs for all the system components and stronger coordination among health stakeholders at both the regional and national levels. In the short term, a document and biorisk mentorship program is needed where hospital-level laboratories mentor, develop specific templates and share with facilities for adaptation. In the medium to long term, an equipment calibration and maintenance program should be implemented across laboratories.

## Data Availability

The data that support the findings of this research will be made available upon reasonable request from the corresponding author.

## Acknowledgement

We are grateful to the Ghana Health Service for providing the initial approval to conduct this research. We also thank the Regional Director of Health Service, District Directors and Medical Superintendents of the Savannah Region for granting permission to collect the data in the region for this study.

## Supporting information

**S1 Fig. Map of Savannah Region showing location of selected study sites**

**S2 Fig. Distribution of median percentage score by laboratory module indicator (N=10)**

**S3 Fig. Median gap score for laboratory needs from smallest (0) to biggest (5), Savannah Region, 2025**

**S4 Fig. Proportion of monthly test done per discipline, Savannah Region, Ghana, 2025**

**S1 Table. Characteristics of laboratories in the WHO LAT assessment, Savannah Region, Ghana, 2025**

**S2 Table. Median assessment score by laboratory type in the Savannah Region, Ghana, 2025**

